# Development, validation and clinical utility of Short-term AdVerse-effects of Electroconvulsive therapy (SAVE) Checklist

**DOI:** 10.1101/2024.01.24.24301691

**Authors:** Chithra Uppinkudru, Harsh Pathak, K Raj Kumar, S Bridgit, Kiran Bagali, Makarand Pantoji, Nathiya Ezhumalai, Rujuta Parlikar, Vyoma Shah, Srinivas Balachander, Vanteemar S Sreeraj, Urvakhsh Meherwan Mehta, Preeti Sinha, Shyam Sundar Arumugham, Ganesan Venkatasubramanian, Jagadisha Thirthalli

## Abstract

Electroconvulsive therapy (ECT) is one of the most effective treatments in psychiatry. However, it has many cognitive and non-cognitive adverse effects (AEs). There are lacunae in the systematic literature on non-cognitive AEs that need a standard, comprehensive and specific clinical tool to evaluate this. Hence, a checklist of short-term AEs of ECT (SAVE) with a 2-phase assessment was developed. Content validation was done using 15 experts’ ratings and predefined content validity ratio and index (CVR and CVI) in a two-stage modified Delphi method. The checklist had a good CVR and CVI with a final tool of 39 items. The tool was sensitive and identified the non-cognitive AEs after ECT. Cardiovascular and musculoskeletal systems displayed the highest incidence. Many participants exhibited delayed recovery in orientation, gait, and stance, highlighting a necessity for meticulous monitoring. SAVE is the first standardised tool to assess short-term ECT-related AEs systematically. This checklist identified a clinically significant incidence of AEs that would otherwise remain unnoticed. Its regular use may enhance the safety of ECT and patient comfort by supporting early identification and intervention for AEs. However, given the transient nature of AEs, further studies are needed to determine their predictive validity for long-term consequences.

## Introduction

Electroconvulsive therapy (ECT) has been an effective treatment in managing several psychiatric conditions with a much quicker response than medications. It is a potentially lifesaving treatment for those having catatonia, neuroleptic malignant syndrome, and severe suicidal ideations (Marks, 1984). The use of anaesthesia and seizure induction have physiological effects on the cardiovascular system (CVS), central nervous system (CNS), and musculoskeletal systems. With such multiple sessions in the course of ECT, one can expect adverse effects (AEs), and some may persist after the treatment has been completed (Sackeim et al., 2007).

The occurrence of a range of adverse events (AEs) associated with ECT has been reported across a range of <1% - 85%, and common side effects like headache and myalgia are typically characterized as mild and temporary. The presence of concurrent medical comorbidities is recognized to exacerbate the incidence of these side effects (Andrade et al., 2016; Zielinski et al., 1993). This knowledge has come predominantly from retrospective studies that used non-standardized assessments. Unlike the cognitive AEs related to ECT, the non-cognitive AEs are not studied systematically (Bassa et al., 2021; Lisanby et al., 2022). The development of structured validated tool to measure the AEs in clinical or research scenarios will help in their better identification, quantification, and management, thereby improving the standard of practice. It may also reduce morbidity and mortality associated with ECT. The current study aimed to develop and validate a checklist for assessing short-term (<48 hours) AEs related to ECT. This study also assessed the feasibility and the utility of the checklist in routine clinical practice.

## Methods

This study was conducted at the National Institute of Mental Health and Neuro Sciences (NIMHANS), a tertiary care center in India, from June 2022 to August 2023. The study protocol was approved by the Institutional Review Board and Ethics Committee [IRB Min. No. NIMHANS/36^th^ IEC (BEH.SC.DIV.)/2022)]. Appropriate written informed consent was obtained from experts and patients undergoing ECT treatment for the respective phases of the study. Caregivers provided consent when the patients did not have the capacity to consent.

### ECT Procedure

At our center, the standard ECT procedure involves the use of bifrontal (BF) ECT montage for induction of seizure, with thiopentone administered at a dosage of 2-4 mg/kg body weight for general anesthesia and succinylcholine at a dosage of 0.5-1 mg/kg body weight as a muscle relaxant. Vital signs (including heart rate (HR), blood pressure (BP), respiratory rate (RR), and oxygen saturation (SpO_2_)) are monitored at baseline, after the administration of the ECT charge, and before the patient is discharged from the ECT suite. Deviation from the above protocol is usually based on clinical conditions/needs like medical comorbidities and the observed adverse effects.

#### Part 1: Development and content validation

The development and validation of the Short-term AdVerse-effects of ECT (SAVE) Checklist were conducted as described below. Figure 1 briefly outlines the development and validation phases along with assessing the utility of SAVE.

**Figure 1.**
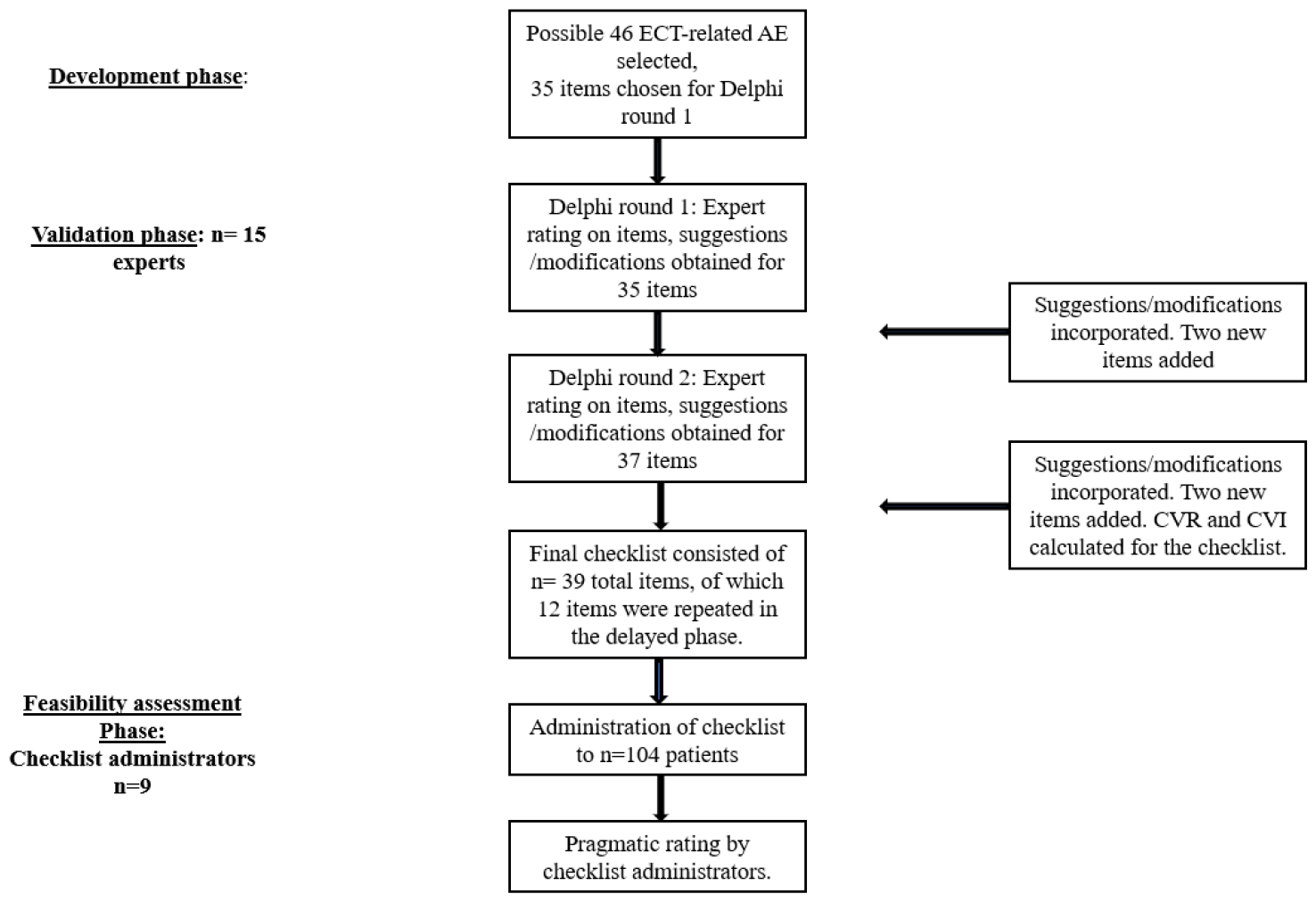
Flowchart of the study methodology. AE-Adverse effects, CVR-Content Validity Ratio, CVI-Content Validity Index, ECT-Electroconvulsive therapy

##### 1. Tool Development

A literature review was conducted to identify the reported short-term AEs of modified ECT. We considered those AEs that developed within 48 hours of an ECT session. This was based on the practice of administering the next ECT session after 48-72 hours. After a detailed discussion among the panel of authors, 35 AEs from the list of 46 AEs were finally selected for validation. The frequency and utility of an AE and the ability to assess it were the factors considered for selection. Two time periods for assessment were defined as “immediate period” referring to the beginning of the ECT session till the immediate recovery from the treatment procedure, and “delayed period” referring to the period between shifting the patient to the ward till about 48 hours of the treatment procedure. Some items were represented in both the immediate and delayed assessment phases based on their possible occurrence during these two phases. An operationalised definition was provided for each item in the list to enhance the objectivity of the rating. The items were organised based on the organ systems (e.g., Cardiovascular, central nervous system, etc.) and the phases of assessment (immediate and delayed).

##### 2. Content Validation

Content validity was assessed by the extent to which expert members of a Content Evaluation Panel perceive overlap between the test and the required job performance domain. The selected items were validated through expert input using the two-stage modified Delphi method (Clayton, 1997). To validate the tool, we approached 15 experts, an adequate number to reach saturation for this method (Clayton, 1997). The experts consisted of consultant psychiatrists in Adult (n=9) as well as Child and Adolescent Psychiatry (n=1), staff nurses in ECT services as well as general psychiatry wards (n=2), consultant anesthetists (n=2), and psychiatry senior resident (n=1). They all had experience prescribing, administering and/or monitoring ECT in at least 100 patients. Experts rated each item for their appropriateness and clarity of writing on a 7-point Likert scale (1-strongly disagree; 2: disagree; 3: somewhat disagree; 4: neither agree nor disagree; 5: somewhat agree; 6: agree; and 7: strongly agree). Further, they rated the entire checklist for its completeness, organisation, and clarity of purpose using the same Likert scale. We also requested them to suggest any additional relevant items for the checklist.

A second version of the checklist was created by incorporating the new items and modifications recommended by the experts. This was again rated by the experts using the Likert scale. The panel of authors finalised the items based on evaluating experts’ comments and results of content validity ratio/index (CVR/CVI, as described in the section on statistical analysis).

#### Part 2: Utility of the checklist

##### 1. Feasibility

Patients (>18 years) receiving ECTs were assessed using the developed checklist. The AE checklist was administered by nine trained assessors from different mental health and allied backgrounds, including psychiatrists (n=5), psychologists (n=1), psychiatric staff nurses (n=1), and psychiatric social workers (n=2). All of them were trained to administer the checklist to build consensus on the interpretation of the manual before the initiation of the study. The physical examination aspects of the checklist were always done by a psychiatrist/nurse. After the completion of data collection, the assessors rated the ease of administration, clarity of definitions, and procedures over a 10-point Likert scale, along with the average time taken for administration in each phase.

##### 2. Incidence of AE

While psychometrically validating a clinical tool, collecting the data in a sample size equal to 2 to 20 per item is recommended as optimal (Anthoine et al., 2014). This required a minimum of 60 to 600 samples for 30 independent items in the checklist. In our center, about 15 new patients are initiated on ECT weekly, receiving an average of about 6 ECT sessions each. Considering the above and accounting for any dropouts/missing data, our study focused on assessing a minimum of 100 consecutive patients (>18 years of age) undergoing ECT for various psychiatric indications for all the sessions during the course of ECT (∼600 ECT sessions). Patients’ short-term AEs were recorded throughout the course of their ECT by the above-mentioned nine assessors.

## Statistical Analysis

The ratings provided by the experts were utilised to compute the CVR and CVI. CVR is an item-level statistic, and it was calculated using the formula CVR = (Ne - N/2) / (N/2), where Ne represents the number of panellists rating the item as “essential” (i.e., the number of panellists who scored >5 on the Likert scale), and N stands for the total number of expert panellists. Essentiality in this study refers to the five different aspects of the checklist that were rated by experts, namely, “appropriateness of the item”, “clarity of writing of the item”, “organisation of the checklist”, “clarity of purpose of the checklist”, and “completeness of the checklist”. Based on Lawshe’s table, a CVR of 0.49 is considered a critical value in a study with 15 experts (Gilbert and Prion, 2016). A score higher than the critical value implied that the agreement among the experts was not by chance and the lesser ones needs consideration for removal/major modification. CVI was calculated by averaging the CVR scores. Values closer to 1 are considered a good measure of the overall content validity of the tool. Further, descriptive statistics were applied to calculate the incidence of AEs and utility.

## Results

### Part 1: Development and validation

#### Content validation

In the first round, experts rated 35 items at the item and checklist levels (Supplementary Table 1). All the items and checklist-level domains cleared the pre-defined threshold of CVR with CVI>0.9 across all 5 aspects. However, suggestions like introducing new items, modifying definitions, and rephrasing AE items were considered and revised with consensus among the panel of authors. For the second round of the Delphi process, the SAVE checklist had 37 items. In this round, the CVI for all 5 aspects was found to be ≥0.94 (Figure 2), showing an overall high content validity of the tool. However, CVR for clarity of definitions for “Significant Tachycardia’’ and “Significant Bradycardia” was less than 0.49 even though the appropriateness had a score of 1. Hence, their definitions were modified addressing the concerns remarked by the experts. (See Supplementary for the evolution of the checklist items and Supplementary Table 1 and Supplementary Figures 1A and 1B for detailed ratings).

**Figure 2:**
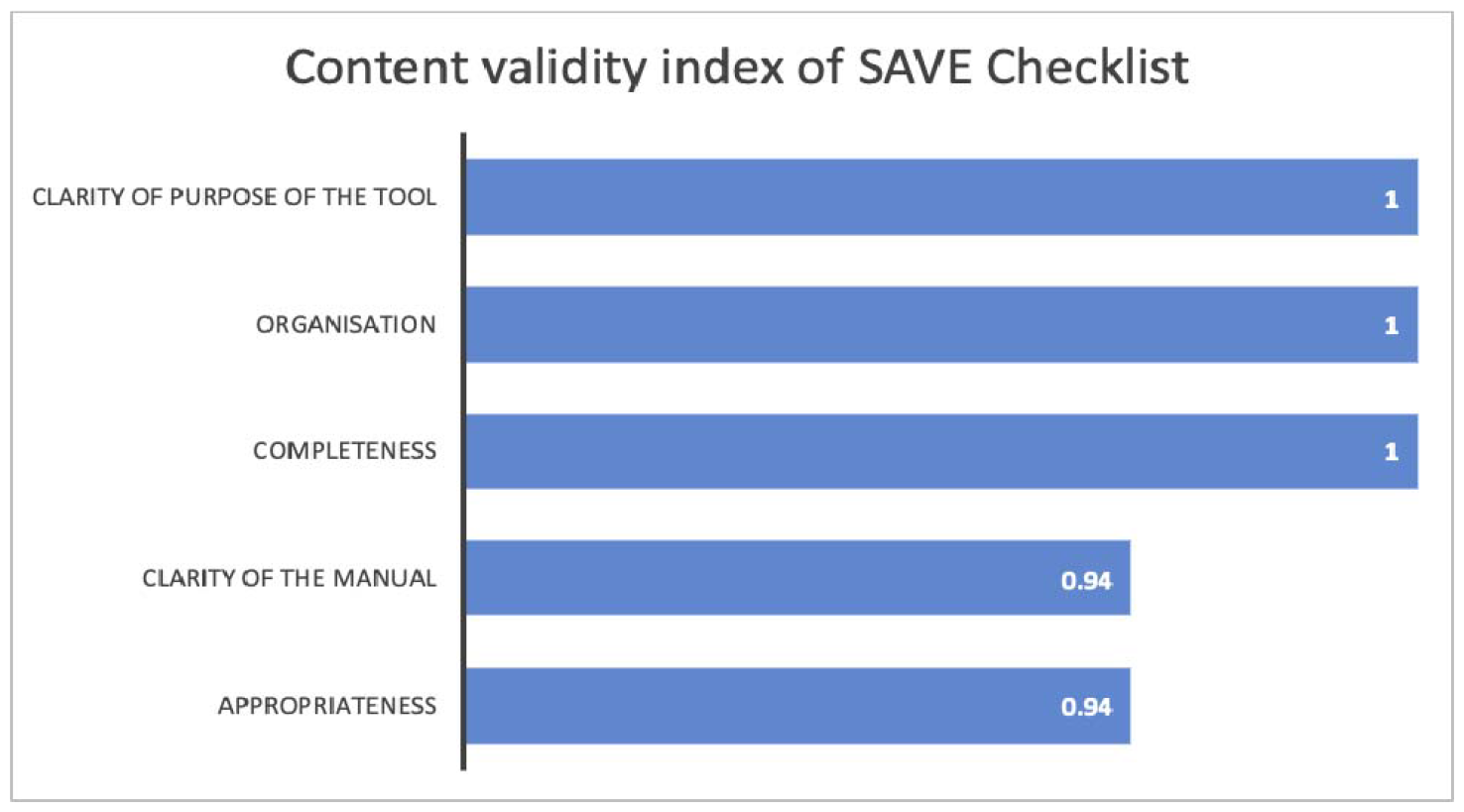
Content validity index of SAVE.

Following recommendations from experts, the final checklist comprising 39 items, of which 12 were repeat items in the delayed assessment phase, was used for the subsequent part of the study. (This free-to-use checklist can be accessed at https://www.neuromodulation-crc.ipsych.in/clinical-tools/)

### Part 2: Assessment of incidence of AEs

The tool was administered to 104 patients, 49 (47%) females. The average age of participants was 37.1 ± 14.6 years. Around 56% of the participants were in the age group of 26-44 years (see Supplementary Tables 2 and 3). A total of 802 ECT sessions were administered in these patients, with each individual undergoing an average of 7.71 (±3.85) sessions. However, due to logistic reasons, the immediate AEs checklist was administered for 764 ECT sessions, whereas delayed AEs could be administered for 647 ECT sessions. Bifrontal ECT was administered to 96 (92.3%) of the participants.

The nine trained assessors who administered the checklist further validated the procedure of the tools. Overall, they rated the checklist as easy to administer, with clear definitions of each item and clarity in the procedure. Most reported about 15 minutes as the additional time needed for the immediate phase of assessments and 10 minutes for the delayed phase of assessments (Table 1).

**Table 1:** Feasibility and utility of the checklist.

| Measures | Mean ± SD | Range |
| --- | --- | --- |
| Ease of Administration<br>(1=Very Difficult 10= Very Easy) | $8.67 \pm 0.71$ | 8-10 |
| Clarity of Definition<br>(1=Entirely Unclear 10= Very Clear) | $8.33 \pm 0.71$ | 7-9 |
| Clarity of the Procedure<br>(1=Entirely Unclear 10= Very Clear) | $8.22 \pm 0.67$ | 7-9 |
| Immediate Assessment- Time Required (In Minutes) | $16.11 \pm 3.33$ | 15-20 |
| Delayed Assessment- Time Required (In Minutes) | $7.78 \pm 2.64$ | 5-10 |

#### Adverse effects

##### Cardiovascular system

Hypertension was the commonest cardiovascular effect noted in this study, with nearly 90% (n=94) had elevated BP soon after the ECT-induced seizures, where 35.57% (n=37) participants had persisting elevation of BP during the early recovery period. Hypotension and desaturation were noted in 40.38% and 37.5% of patients after ECT, respectively. Incidences of hypotension increased from post-ECT to recovery period from 8.25% to 18.98% of sessions, but incidences of desaturation were reduced from 8.77% to 3.01% of sessions. Other issues like arrhythmias or asystole were not observed in the sessions (Table 2). Most of the cardiovascular adverse effects, except tachycardia and hypertension, happened in a median of 1-2 sessions during the course of ECT.

**Table 2:** Adverse effects - patient and session wise, their percentage, range of occurrence among patients receiving treatment.

| Adverse effect (AE) | Number of patients (N=104) | Percentage of patients | Number of sessions | Percentage of sessions* | Range of occurrences of AE across sessions (median) |
| --- | --- | --- | --- | --- | --- |
| **A. Immediate Assessments for Adverse effects (number of sessions- 764)** |  |  |  |  |  |
| **1. Cardiovascular System** |  |  |  |  |  |
| a. Significant Tachycardia |  |  |  |  |  |
| Baseline | 5 | 4.80 % | 10 | 1.13 % | 1 to 5 (1) |
| After ECT | 59 | 56.73 % | 147 | 19.2 % | 1 to 10 (2) |
| Recovery Period | 43 | 41.34 % | 88 | 11.5 % | 1 to 7 (1) |
| b. Significant Bradycardia |  |  |  |  |  |
| Baseline | 4 | 3.85 % | 5 | 0.7 % | 1 to 2 (1) |
| After ECT | 6 | 5.77 % | 7 | 0.9 % | 1 to 2 (1) |
| c. Significant Hypertension |  |  |  |  |  |
| Baseline | 30 | 28.84 % | 56 | 7.33 % | 1 to 8 (1) |
| After ECT | 94 | 90.38 % | 330 | 43.19 % | 1 to 11 (3) |
| Recovery Period | 37 | 35.57 % | 66 | 8.64 % | 1 to 4 (1) |
| d. Significant Hypotension |  |  |  |  |  |
| After ECT | 42 | 40.38 % | 67 | 8.25 % | 1 to 5 (1) |
| Recovery Period | 68 | 65.38 % | 145 | 18.98 % | 1 to 7 (2) |
| e. Significant Desaturation |  |  |  |  |  |
| After ECT | 39 | 37.5 % | 67 | 8.77 % | 1 to 7 (1) |
| Recovery Period | 12 | 11.54 % | 23 | 3.01 % | 1 to 4 (1.5) |
| f. Asystole | 0 | 0 % | 0 | 0 % | 0 |
| g. Arrhythmia | 0 | 0 % | 0 | 0 % | 0 |
| **2. Gastrointestinal System** |  |  |  |  |  |
| a. Oral Injuries | 4 | 3.80 % | 4 | 0.52 % | 1 (1) |
| b. Loosening/Breaking of Teeth | 3 | 2.89 % | 3 | 0.39 % | 1 (1) |
| **3. Respiratory System** |  |  |  |  |  |
| a. Prolonged Apnoea | 1 | 0.97 % | 1 | 0.13 % | 1 (1) |
| b. Suspected Aspiration | 1 | 0.97 % | 1 | 0.13 % | 1 (1) |
| **4. Central Nervous System** |  |  |  |  |  |
| a. Delirium | 85 | 81.73 % | 223 | 29.18 % | 1 to 14 (2) |
| b. Prolonged Time For Orientation | 79 | 75.96 % | 197 | 25.78 % | 1 to 8 (2) |
| c. Prolonged Seizure | 2 | 1.90 % | 2 | 0.26 % | 1 (1) |
| **5. Musculoskeletal System** |  |  |  |  |  |
| a. Fall | 0 | 0 % | 0 | 0 % | 0 |
| b. Suspected Fracture/ Dislocation | 0 | 0 % | 0 | 0 % | 0 |
| c. Prolonged Gait Abnormality | 82 | 78.85 % | 208 | 27.22 % | 1 to 15 (2) |
| d. Prolonged Stance Abnormality | 33 | 31.73 % | 167 | 21.85 % | 1 to 15 (2) |
| **6. Genitourinary System** |  |  |  |  |  |
| a. Enuresis | 5 | 4.80 % | 6 | 0.78 % | 1 to 2 (1) |
| b. Encopresis | 1 | 0.97 % | 1 | 0.13 % | 1 (1) |
| **7. Other Adverse-Effects** |  |  |  |  |  |
| a. Fever | 0 | 0 % | 0 | 0 % | 0 |
| b. Delayed onset seizure | 0 | 0 % | 0 | 0 % | 0 |
| c. Tardive seizure | 0 | 0 % | 0 | 0 % | 0 |
| **B. Delayed Assessment For Adverse-Effects (number of sessions- 647)** |  |  |  |  |  |
| **1. Gastrointestinal System** |  |  |  |  |  |
| a. Oral Injury | 4 | 3.84 % | 4 | 0.62 % | 1 (1) |
| b. Loose Tooth | 0 | 0 % | 0 | 0 % | 0 |
| c. Vomiting | 13 | 12.5 % | 21 | 3.24 % | 1 to 5 (1) |
| d. Nausea | 23 | 22.12 % | 31 | 4.79 % | 1 to 3 (1) |
| **2. Musculoskeletal System** |  |  |  |  |  |
| a. Fall | 7 | 6.70 % | 7 | 1.08 % | 1 (1) |
| b. Myalgia | 33 | 31.73 % | 48 | 7.42 % | 1 to 3 (1) |
| c. Headache | 62 | 59.60 % | 171 | 26.42 % | 1 to 12 (2) |
| d. Temporomandibular pain | 29 | 27.90 % | 36 | 5.56 % | 1 to 3 (1) |
| **3. Central Nervous System** |  |  |  |  |  |
| a. Awareness under anaesthesia | 3 | 2.89 % | 3 | 0.46 % | 1 (1) |
| b.Sedation | 23 | 22.11 % | 28 | 4.38 % | 1 to 2 (1) |
| c.Delirium | 5 | 4.80 % | 5 | 0.77 % | 1 (1) |
| d.Subjective cognitive impairment | 38 | 36.53 % | 57 | 8.80 % | 1 to 3 (1) |
| **4. Other Adverse-Effects** |  |  |  |  |  |
| a.Fever | 5 | 4.80 % | 7 | 1.08 % | 1 to 2 (1) |
| b.Phlebitis | 7 | 6.73 % | 7 | 1.08 % | 1 (1) |
| c.Todd phenomenon | 0 | 0 % | 0 | 0 % | 0 |
| d.Tardive seizures | 1 | 0.96 % | 1 | 0.15 % | 1 (1) |

##### Central Nervous system

About 82% (n=85) of participants had delirium in the form of agitation (n=6) or delayed time for orientation (n=79) during the immediate assessment period. Thus, the hypoactive type of delirium was the most common to be observed. It should be noted that orientation could not be systematically assessed in about 63 (8.2%) patient sessions due to non-cooperation owing to underlying psychiatric illness (Figure 3). Prolonged seizure was observed in two patient sessions (146s and 185s), which were promptly aborted by additional doses of benzodiazepines. In the delayed assessment of AEs, the proportion of patients reporting subjective cognitive impairment was 36.53% (n=38). About 4.8% (n=5) of the participants were thought to have delirium in the form of disorientation (n=1) or agitation (n=4). Sedation was reported in 4.38% of patient sessions. Delay in orientation, sedation, and headache tended to recur in the same participant more often than not. Awareness under anesthesia was described by three different patients (0.46%) in one session each.

**Figure 3:**
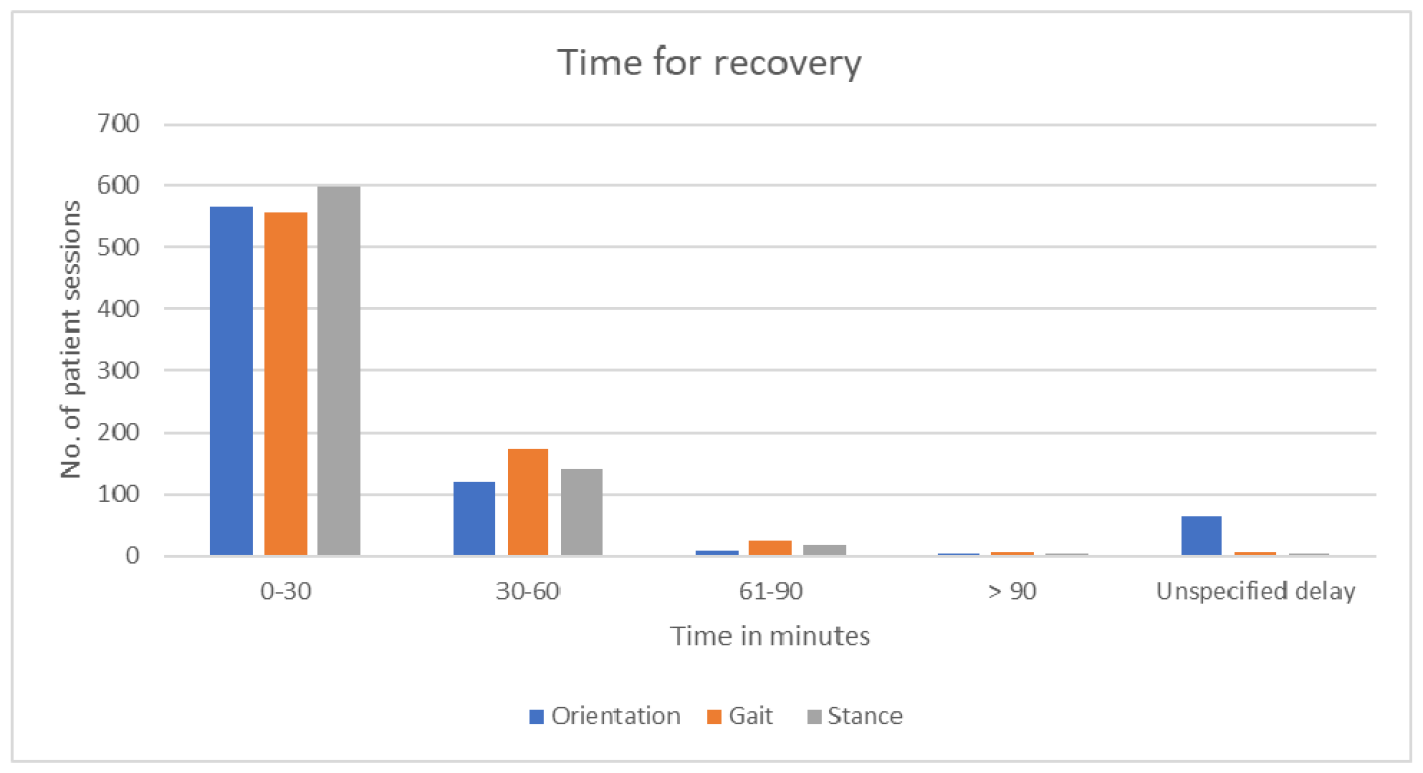
Gait, stance, and orientation recovery time measured across patient ECT sessions. Unspecified delay-refers to patients whose time-for-recovery could not be assessed

##### Musculoskeletal system

Gait abnormality (Figure 3) was seen in 78.85% (n=82) of the participants, and prolonged time for stance recovery was seen in 31.73% (n=33) of patients. Temporomandibular (TM) pain (n=36, 5.56%) and myalgia (n=48, 7.42%) were commonly reported in patient sessions. In the delayed assessment period, a fall was reported for 7 independent patient sessions (1.08%) and was observed to be a non-recurring AE. However, no major musculoskeletal injuries were noted secondary to falls.

##### Respiratory system

There were two respiratory AEs in the immediate assessment period (0.13%) in the form of prolonged apnoea and aspiration.

##### Gastrointestinal system

Nausea and vomiting were the commonly reported delayed gastrointestinal AEs. A broken tooth was noted in 4 patients.

##### Genitourinary system

Enuresis immediately after ECT was observed in 6 sessions (0.78%). Encopresis was observed in one patient session.

##### Other rare AEs

There was one possible report of tardive seizure during the delayed assessment of AEs. Other AEs include shivering (n=2, 0.26% patient sessions), dizziness (n=8, 1.04% patient sessions) in the immediate period, and phlebitis (n=7, 1.08% patient sessions), fever (n=7, 1.08% patient sessions) during the delayed assessment period.

## Discussion

The study introduced a novel checklist for evaluating short-term AEs of ECT (SAVE) designed to serve as a systematic screening tool for short-term AEs. It provides an objective and comprehensive assessment of the range of known AEs.

Developing the checklist involving experts working closely with the patients receiving ECT and greater agreement among these experts is its strength. Further, feasibility was evaluated in a reasonably large sample adequate to identify common AEs. This checklist offers a systematic assessment of unreported and understudied AEs and would assist in enhancing the safety of the ECT in clinical and research settings. We recommend using this alongside tools to assess ECT-related cognitive AEs such as Battery for ECT Related Cognitive Deficits (B4ECT ReCoDe) (Viswanath et al., 2013) or ElectroConvulsive Therapy Cognitive Assessment scale (ECCA) (Hermida et al., 2020).

### Incidence of side effects

A clinically significant proportion of individuals were observed to experience adverse events (AEs) across various physiological systems, and these were predominantly transient and mild, consistent with findings from the literature (Melzer-Ribeiro et al., 2023; Rhee et al., 2022). AEs related to the cardiovascular and nervous systems were commonly noted during the immediate phase. Previous research has indicated subjective memory impairment ranging from 26-75% (Kheirabadi et al., 2020; Zhang et al., 2022), hypertension in 55.4% (Antosik-Wójcinska et al., 2022), and orientation abnormalities in 18-33% (Antosik-Wójcińska et al., 2022; Kheirabadi et al., 2019). The reported incidence in our study is higher than those recorded in earlier studies that were either based on patients’ self-report (Ekstrand et al., 2022) or retrospective review of charts(Antosik-Wójcińska et al., 2022). This was especially true for side effects like significant hypertension, delirium, significant desaturation, gait, and stance abnormality. In a prospective study (n=90) examining AEs immediately post-treatment and after 24 hours, confusion, headache, myalgia, amnesia, and vertigo were reported at rates of 24.4%, 80%, 53.3%, 28.9%, and 24.4%, respectively. Persistent symptoms after 24 hours included amnesia (5.5%), vertigo (5.5%), headache (16.67%), and myalgia (28.8%) (Ekstrand et al., 2022). Consistent with prior research, headache, TM pain, and myalgia were commonly observed in nearly 50% of the patients despite receiving modified ECTs (Andrade et al., 2016; Zhang et al., 2022; Kheirabadi et al., 2019). The practice of using relatively lower doses of muscle relaxants could be a contributing factor. But, succinylcholine could also induce myalgia, and its contribution needs systematic evaluation. Conversely, gastrointestinal and genitourinary AEs were reported less frequently. Other effects like post-ECT shivering, phlebitis, and mild sedation seem to be contributed by anesthesia-related aspects of ECT.

### Practical utility in routine clinical practice

The checklist appears to be designed for easy clinical application, with an average evaluation time of 16 minutes for immediate AEs and 8 minutes for delayed assessments. Any trained single administrator could apply the checklist with minimal assistance from the psychiatrist, anesthetist, and/or ECT-administering nurses. Measures like asystole, arrhythmia and prolonged seizure may be better identified using ECG and EEG and require an expert to monitor and interpret, which is the case in routine ECT practice. Barring these items, other checklist items can be collected by any minimally trained mental health professional.

Systematic assessment of pre-ECT orientation, gait, and stance is difficult in certain patients who are too symptomatic and non-cooperative (conditions like agitation and catatonia). Application of clinical knowledge would be needed in such cases to determine the post-ECT recovery. Modifying the evaluation can involve more focused observation of general behaviour and speech, asking simple questions about the patient’s name and residence, and asking them to follow some basic commands (Martin et al., 2018). The checklist’s organization based on physiological systems and differentiation between immediate and delayed phases enhances its clinical utility. Two-time assessments minimize the risk of overlooking AEs, ensuring a thorough evaluation. The objective design reduces potential errors and subjectivity, emphasizing its applicability.

A transient rise in blood pressure is an expected physiological response during ECT. This finding, seen in nearly every individual undergoing ECT, may raise questions about the clinical significance of defining it as an adverse effect. We recommend that clinicians/researchers adopting this checklist may have to reconsider the cut-off or incorporate the persistence of side effects beyond a designated time frame to define it as significant hypertension. Further studies could focus on identifying the optimal definition that can have good predictive validity for long-term consequences. The checklist also systematically assessed the time to orientation, stance, and gait; the latter two AEs used a modification of the SARA scale for assessment (Grobe-Einsler et al., 2023). These events are often neglected in regular clinical settings and might be relevant in avoiding complications. Previous studies have indicated a connection between prolonged orientation time and cognitive deficits; therefore, assessment during the immediate ECT period has an implication in patient outcomes (Martin et al., 2018). Their clinical implication on longer-term cognitive AEs needs further evaluation. The study also highlights that many individuals required more than 30 min for their gait and stance to return to baseline level. This finding holds the potential for improving the standards of post-ECT patient care in recovery rooms. This may largely prevent many consequent AEs, like falls and cardiovascular instabilities. A higher incidence of some of these side effects was noted during the earlier sessions.

When AEs were noted, it was reported to the in-charge treating team to prevent their recurrence through modification of the dose of the anesthetic agent /muscle relaxant or ongoing pharmacotherapy along with other precautions and application of risk mitigation strategies. This could have influenced the incidence rates estimated in this study. Nevertheless, this suggests that regular monitoring and reporting to the clinical/ECT administration team can help take proper corrective and precautionary steps to enhance the safety of ECT.

Presence of awareness under anesthesia in many participants, where they persisted in having a vague memory of the muscle relaxant-induced apnea. This event would mostly remain unreported unless specifically checked for and could lead to phobic anxiety towards the ECT procedure. This could impact patient attitude and adherence toward continuing the treatment. Reassurance, ascertaining the level of consciousness, increasing anesthesia dose, and delaying the administration of muscle relaxants were considered when this AE was noted.

When considering the clinical applicability of the scale, several unresolved factors persist: If AEs are transient and mild, can the pre-anesthetic standards be more lenient? Additionally, determining the optimal timing for intervention in addressing these side effects, understanding the long-term repercussions of repeated anesthesia induction and stimulation of the autonomic system, exploring trajectories of the observed AEs during the course of ECT, and assessing whether intervention strategies should be tailored for individuals with medical comorbidities are essential considerations. It was also observed that the decision to intervene to handle these transient AEs (e.g., hyper/hypotension) was subjective and varied across clinicians (psychiatrists/anaesthesiologists). This points to the need for further work on systematically identifying the AEs and consolidating standards of care involving all stakeholders like psychiatrists, anesthesiologists, nurses, and patients.

The study and the checklist have certain limitations. A notable number of patients were reported to have delirium/agitation. But, in a good subset of the patients, reorientation could not be ascertained due to the underlying clinical condition. The study had no and limited representation of the child/adolescent and geriatric patient populations, respectively. The study could not establish causality of symptoms, which falls outside the scope of the present paper, but this work provides a promising platform to explore these avenues in conjunction with safe ECT practice in the future. Future studies using this tool in special population groups would further enhance the safer practice of ECT. Since this study was powered to evaluate common and very common side effects of ECT, comments on the reliability of other findings related to side effects cannot be made.

## Conclusions

AEs related to ECT have important implications for therapeutic outcomes and treatment acceptance in patients and their caregivers. So far, systematic tools have existed only for assessing cognitive adverse effects. This comprehensive checklist has immense potential to facilitate better evaluation and management of other AEs and enhance patient care.

## Data Availability

All data produced in the present study are available upon reasonable request to the authors.

https://www.neuromodulation-crc.ipsych.in/clinical-tools/

## Acknowledgement

CU, HP, RKK, BS, KB, MP, NE, and RP are supported by the Department of Biotechnology (DBT)-Wellcome Trust India Alliance (IA/CRC/19/1/610005). VS is supported by Rohini Nilekani Philanthropies (G-202303-00523) funded Centre for Brain and Mind (CBM). VSS acknowledges the support of the India-Korea joint program cooperation of science and technology by the National Research Foundation (NRF) Korea (2020K1A3A1A68093469), the Ministry of Science and ICT (MSIT) Korea, and the Department of Biotechnology (India) (DBT/IC-12031(22)-ICD-DBT). SSA acknowledges support from Department of Biotechnology (DBT) – Wellcome Trust India Alliance (IA/CPHI/18/1/50393). GV acknowledges the support of the Department of Biotechnology, Government of India (BT/HRD-NBA-NWB/38/2019-20(6)).

