## Supplement File for "Development, validation and clinical utility of Short-term AdVerse-effects of Electroconvulsive therapy (SAVE) Checklist"

**Supplementary material:**

**Details of the evolution of checklist items**: After the first round of Delphi, four items on heart rate and blood pressure were modified (Highest/lowest heart rate and highest/lowest blood pressure to significant tachycardia/bradycardia and significant hypertension/hypotension, respectively). Phlebitis and hypoactive delirium were added. After the 2nd round of Delphi, agreement on the clarity of operational definition of the tachycardia and bradycardia was poor, with CVR less than the preset threshold of 0.49 (0.46 each), though the score for appropriateness had full agreement(CVR=1). After considering the remarks from the experts, the operational criteria were modified. Additionally, one new item was incorporated under the CVS assessment: "significant desaturation," defined as SpO_2_ reading below 90% during observation periods. Also, two items were introduced in the musculoskeletal system assessment: "prolonged time for gait recovery" and "prolonged time for stance recovery," defined as abnormalities in gait and stance, respectively, lasting more than 30 minutes. These two items were evaluated using the Scale for the Assessment and Rating of Ataxia (SARA) scale (Grobe-Einsler et al., 2023). Furthermore, based on expert suggestions, two existing items (hypoactive and hyperactive delirium) were merged to enhance clarity and efficiency.

The finalised tool had two phases for assessment: The immediate phase and the delayed phase. The immediate phase involved assessments during the pre-ECT preparation, administration of ECT, and post-ECT recovery until the patient was shifted back to the ward. The delayed phase started from this point till 48 hours after receiving ECT. In our study, the delayed assessment was conducted approximately 24 hours after the last ECT. The checklist was further divided into individual systems (e.g., cardiovascular, gastrointestinal, etc.) for further clarity. AEs that occur during both immediate and delayed phases or persist from the immediate to the delayed phase were mentioned twice in the checklist.

**Supplementary Table 1: Content validity ratio (CVR) of Delphi round one and two of checklist validation at item level and checklist level**

| **Checklist Items** | **CVR (Round 1)** | | **CVR (Round 2)** | |
| --- | --- | --- | --- | --- |
|  | **A** | **C** | **A** | **C** |
| Highest Heart Rate | 0.6 | 0.73 | - | - |
| Significant Tachycardia | - | - | 0.87 | 0.46 |
| Lowest Heart Rate | 0.6 | 0.73 | - | - |
| Significant Bradycardia | - | - | 0.73 | 0.46 |
| Highest Blood Pressure | 0.6 | 0.73 | - | - |
| Significant Hypertension | - | - | 1.00 | 0.87 |
| Lowest Blood Pressure | 0.6 | 0.73 | - | - |
| Significant Hypotension | - | - | 1.00 | 0.87 |
| Asystole | 0.87 | 0.87 | 0.87 | 1.00 |
| Arrhythmia | 0.87 | 0.87 | 1.00 | 1.00 |
| Oral Injury | 1.00 | 1.00 | 1.00 | 1.00 |
| Loosening of teeth | 0.87 | 0.87 | 1.00 | 1.00 |
| Prolonged apnoea | 1.00 | 0.73 | 1.00 | 0.87 |
| Suspected aspiration | 1.00 | 0.87 | 1.00 | 0.87 |
| Hypoactive delirium | - | - | 1.00 | 1.00 |
| Prolonged orientation time | 1.00 | 1.00 | 1.00 | 1.00 |
| Agitation/ delirium | 1.00 | 0.87 | 1.00 | 1.00 |
| Prolonged seizure | 1.00 | 1.00 | 1.00 | 1.00 |
| Delayed onset seizure | 0.87 | 0.87 | 1.00 | 0.87 |
| Tardive seizure | 0.87 | 1.00 | 0.87 | 1.00 |
| Fall | 1.00 | 1.00 | 1.00 | 1.00 |
| Suspected fracture/dislocation | 0.87 | 1.00 | 1.00 | 1.00 |
| Enuresis | 1.00 | 0.87 | 0.87 | 1.00 |
| Encoperesis | 1.00 | 0.87 | 0.87 | 1.00 |
| Fever | 0.74 | 0.87 | 0.74 | 0.87 |
| Loosening of Teeth | 0.6 | 1.00 | 0.74 | 1.00 |
| Intraoral injury | 0.6 | 1.00 | 0.74 | 1.00 |
| Vomiting | 1.00 | 1.00 | 1.00 | 1.00 |
| Nausea | 1.00 | 1.00 | 1.00 | 1.00 |
| Fall | 0.87 | 1.00 | 0.87 | 1.00 |
| Myalgia | 1.00 | 1.00 | 1.00 | 1.00 |
| Headache | 1.00 | 1.00 | 1.00 | 1.00 |
| TM pain | 1.00 | 1.00 | 1.00 | 1.00 |
| Awareness under anaesthesia | 1.00 | 0.87 | 1.00 | 0.87 |
| Sedation | 0.87 | 0.87 | 1.00 | 1.00 |
| Todd phenomenon | 1.00 | 1.00 | 1.00 | 1.00 |
| Tardive seizures | 1.00 | 1.00 | 1.00 | 1.00 |
| Agitation/delirium | 1.00 | 1.00 | 1.00 | 0.87 |
| Subjective memory loss | 1.00 | 1.00 | 1.00 | 1.00 |
| Fever | 0.73 | 0.87 | 0.73 | 0.87 |
| Phlebitis | - | - | 0.87 | 0.87 |
| **Overall tool level** | | | | |
| Completeness | 0.87 | | 1.00 | |
| Organisation | 0.87 | | 1.00 | |
| Clarity of scale purpose | 0.87 | | 1.00 | |

*A=Appropriateness, C=Clarity of writing, TM- Temporomandibular*

**Supplementary Table 2: Socio-demographic details of participants**

| **Variables** | **Frequency (N=104)** | **Percentage** |
| --- | --- | --- |
| **Age** | | |
| 18-25 | 22 | 21.2 % |
| 26-44 | 58 | 55.8 % |
| 45-59 | 12 | 11.5 % |
| 60-90 | 12 | 11.5 % |
| **Gender** | | |
| Female | 49 | 47.1 % |
| Male | 55 | 52.9 % |
| **Diagnosis** (N= 104) | | |
| **A. Organic mental Disorders** | **11** | **10.58%** |
| 1. Organic catatonic disorder | 3 | 2.88% |
| 2. Organic delusional disorder | 2 | 1.92% |
| 3. Organic mood disorder | 6 | 5.77% |
| **B. Schizophrenia, schizotypal and delusional disorders** | **37** | **35.58%** |
| 1. Schizophrenia | 28 | 26.92% |
| 2. Persistent delusional disorders | 1 | 0.96% |
| 3. Acute and transient psychotic disorders | 1 | 0.96% |
| 4. Schizoaffective disorder | 5 | 4.81% |
| 5. Unspecified nonorganic psychosis | 2 | 1.92% |
| **C. Mood disorders** | **29** | **27.88%** |
| 1. Mania with psychotic symptoms | 1 | 0.96% |
| 2. Bipolar affective disorder   a. hypomania b. mania without psychotic symptoms c. mania with psychotic symptoms d. mild- moderate depression e. severe depression without psychotic symptoms f. severe depression with psychotic symptoms | 1  7  14  2  2  2 | 0.96%  6.73%  13.46%  1.92%  1.92%  1.92% |
| 3. Depressive episode   a. Severe depressive episode without psychotic symptoms b. Severe depressive episode without psychotic symptoms | 4  4 | 3.85%  3.85% |
| 4. Recurrent Depressive Disorder   a. moderate episode with somatic symptoms b. severe episode without psychotic symptoms c. severe episode with psychotic symptoms | 2  9  7 | 1.92%  8.65%  6.73% |
| **D. Emotionally unstable personality disorder** | **1** | **0.96%** |

**Supplementary table 3: Medical comorbidities**

| **Comorbidities** | **Frequency** | **Percentage** |
| --- | --- | --- |
| Hypothyroidism | 17 | 16.35% |
| Seizure disorder | 11 | 10.58% |
| Hypertension | 11 | 10.58% |
| Diabetes Mellitus | 9 | 8.65% |
| Anaemia | 8 | 7.69% |
| Vitamin B12 deficiency | 6 | 5.77% |
| Traumatic brain injury | 2 | 1.92% |
| Hyperthyroidism | 1 | 0.96% |
| Bilateral Sensorineural hearing loss | 1 | 0.96% |
| Disc prolapse | 1 | 0.96% |
| Gilbert syndrome | 1 | 0.96% |
| Interstitial lung disease | 1 | 0.96% |
| Malignant neoplasm of ovary | 1 | 0.96% |
| Parkinson's disease | 1 | 0.96% |
| Pituitary microadenoma | 1 | 0.96% |
| Rheumatoid arthritis | 1 | 0.96% |
| Renal agenesis (left) | 1 | 0.96% |

**Supplementary Fig 1A: Appropriateness at the item level in the second round of Delphi**

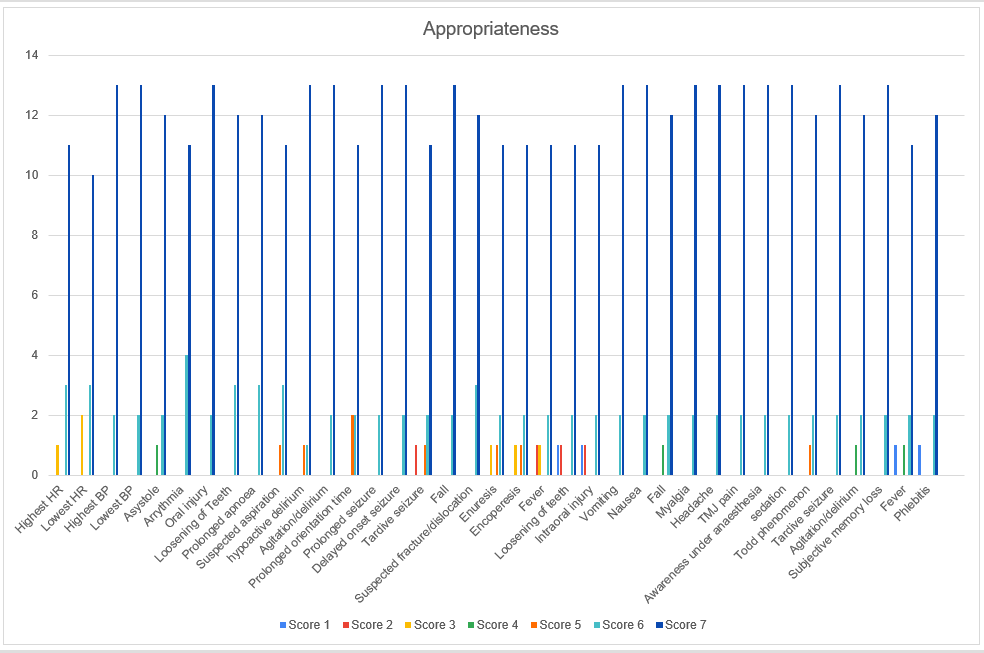

**Supplementary Fig 1B: Clarity at the item level in the second round of Delphi**

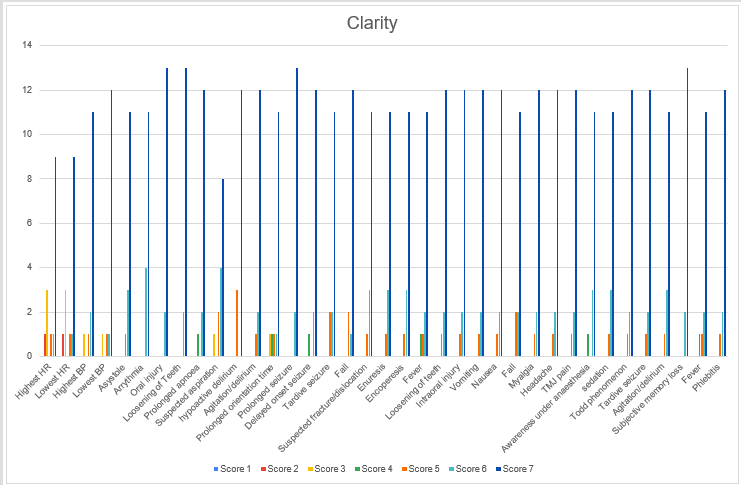
